# UKBAnalytica: an integrated R package for scalable phenotyping and reproducible epidemiological analysis within the UK Biobank Research Analysis Platform

**DOI:** 10.64898/2026.06.19.26356057

**Authors:** Nan He, Ke Mo, Guangchuang Yu, Feiying He

**Author notes:** Corresponding author: Ke Mo, Guangchuang Yu, Feiying He.

## Abstract

UK Biobank provides longitudinal health-related data for approximately 500,000 participants, and its Research Analysis Platform (RAP) has shifted large-scale analyses toward secure cloud-based computation. However, many existing tools address only specific steps of the analytical workflow, leaving a need for an integrated framework that connects multi-source disease phenotyping, survival-ready cohort construction, and downstream analysis on the RAP. Here, we present UKBAnalytica, an extensible R package for scalable phenotyping and integrated analysis of UK Biobank data within the RAP environment. It currently includes 52 predefined baseline variables and a built-in library of 331 curated disease definitions. These definitions are based on multiple UK Biobank data sources, including ICD-10, ICD-9, self-reported conditions, death registry records, algorithmically defined outcomes, and OPCS-4 procedure codes. UKBAnalytica distinguishes prevalent and incident cases, constructs follow-up time, generates analysis-ready survival datasets, and summarizes participant flow. Beyond phenotype construction, UKBAnalytica provides integrated modules for epidemiological analysis, omics analysis, and machine-learning-based modeling and interpretation. By linking endpoint definition with downstream modeling under a consistent data structure, UKBAnalytica reduces repetitive scripting and improves analytical transparency. Furthermore, we demonstrate the package’s practical utility through a case study on chronic obstructive pulmonary disease (COPD) proteomics. The findings align closely with previously reported conclusions, underscoring the robustness and reliability of our analytical framework. This phenotype-centered framework complements existing UK Biobank tools and facilitates reproducible RAP-based biomedical research. UKBAnalytica is freely available at https://github.com/Hinna0818/UKBAnalytica.

## 1. Introduction

UK Biobank has become one of the most influential population-scale biomedical resources for modern health research. By combining deep baseline characterization, longitudinal follow-up, genetic data, lifestyle information, environmental exposures, biomarkers, imaging, and linked electronic health records from approximately 500,000 participants, UK Biobank enables large-scale investigations across epidemiology, genetics, clinical medicine, and multi-omics research^1–3^. This breadth has made UK Biobank a key resource for population-scale studies of disease mechanisms, risk prediction, and precision medicine^4,5^.

However, the same richness that makes UK Biobank powerful also creates substantial analytical challenges. Disease outcomes are rarely derived from a single data field. In practice, researchers often need to integrate hospital inpatient diagnoses, ICD-10 and ICD-9 codes, self-reported conditions, death registry records, algorithmically defined outcomes, and procedure codes such as OPCS-4. These sources must then be aligned with baseline assessment dates to distinguish prevalent from incident cases and to construct valid follow-up time for prospective analyses. As a result, a large proportion of the analytical workload is spent on repetitive data cleaning, source harmonization, endpoint definition, and survival dataset construction. These repetitive steps can reduce analytical efficiency and make it harder to maintain consistent definitions across studies.

Several computational tools have been developed to improve the usability of UK Biobank data. For example, ukbtools reduces the upfront data-wrangling burden by merging UK Biobank files into analysis-ready datasets with descriptive column names, and it further supports disease queries, quality-control summaries, and genetic metadata retrieval^6^. PHESANT was developed to perform automated phenome scans in UK Biobank, enabling systematic association testing across heterogeneous phenotypes^7^. More recently, FastUKB was introduced as an R package-based system with a shiny user interface to simplify UK Biobank RAP data extraction, metadata processing, variable selection, and basic analytical tasks^8^. Other tools have been designed for specific data modalities or RAP-oriented workflow management, such as metabolomics preprocessing or cloud-based variable extraction. Together, these packages have substantially improved the accessibility and efficiency of UK Biobank research.

Despite these advances, a lightweight and phenotype-driven framework for follow-up cohort construction and downstream epidemiological analysis remains needed. Existing tools have substantially improved data wrangling, automated phenome-wide scans, omics-specific preprocessing, and RAP-oriented workflow management, but many practical UK Biobank studies still depend on project-specific scripts to assemble disease endpoints, define baseline exclusions, distinguish prevalent and incident cases, calculate follow-up time, and prepare datasets for downstream modeling. This fragmented workflow may reduce analytical efficiency and create inconsistencies in endpoint definition and time-to-event construction. Therefore, a framework that starts from disease phenotypes and directly supports survival-ready cohort construction and subsequent statistical analysis would provide a useful complement to existing UK Biobank software.

To address these limitations, we developed UKBAnalytica, an extensible R package for phenotype-driven follow-up cohort construction and downstream analysis of UK Biobank RAP data. UKBAnalytica provides a modular framework that connects data preparation, multi-source disease definition, survival-ready cohort construction, epidemiological analysis, omics-oriented analysis, machine learning, and visualization within a consistent data structure. The package supports disease definitions from ICD-10, ICD-9, self-reported conditions, death registry records, algorithmically defined outcomes, and OPCS-4 procedure codes. It further distinguishes prevalent and incident cases, constructs follow-up time, summarizes participant flow, and generates analysis-ready datasets for downstream modeling. The major feature of UKBAnalytica is its endpoint-centered design. By abstracting heterogeneous diagnosis sources into reusable disease definition objects, the package allows researchers to combine multiple evidence sources and apply consistent rules across different diseases. It also provides downstream modules for baseline summaries, Cox regression, subgroup and sensitivity analyses, multiple imputation, propensity score methods, mediation analysis, machine learning, model interpretation, and visualization. These functions can support both conventional epidemiological analyses and biomarker-driven studies involving metabolomic, proteomic, or other high-dimensional data. Overall, UKBAnalytica reduces repetitive scripting and provides a practical framework for more transparent and reproducible UK Biobank research in RAP-based environments.

Finally, we demonstrated the feasibility and utility of UKBAnalytica through a COPD (Chronic Obstructive Pulmonary Disease) proteomics case study conducted within the official UK Biobank RAP environment. Using functions in our package, we defined incident COPD, constructed a survival-ready cohort, processed baseline covariates and plasma proteomic profiles and performed epidemiological analysis. This case study illustrates how UKBAnalytica can connect RAP-derived data with phenotype definition, cohort construction, downstream modeling, and biological interpretation in a single workflow. Together, UKBAnalytica provides a practical and extensible framework for reducing repetitive scripting, improving analytical consistency, and supporting reproducible UK Biobank research.

## 2. Results

### 2.1 The workflow of UKBAnalytica

UKBAnalytica is an integrated workflow package optimized for UK Biobank Research Analysis Platform (RAP) data analysis (Fig. 1). This package provides comprehensive functions for RAP data preprocessing, phenotype definition, and downstream statistical analysis (Table 1).

**Fig. 1.**
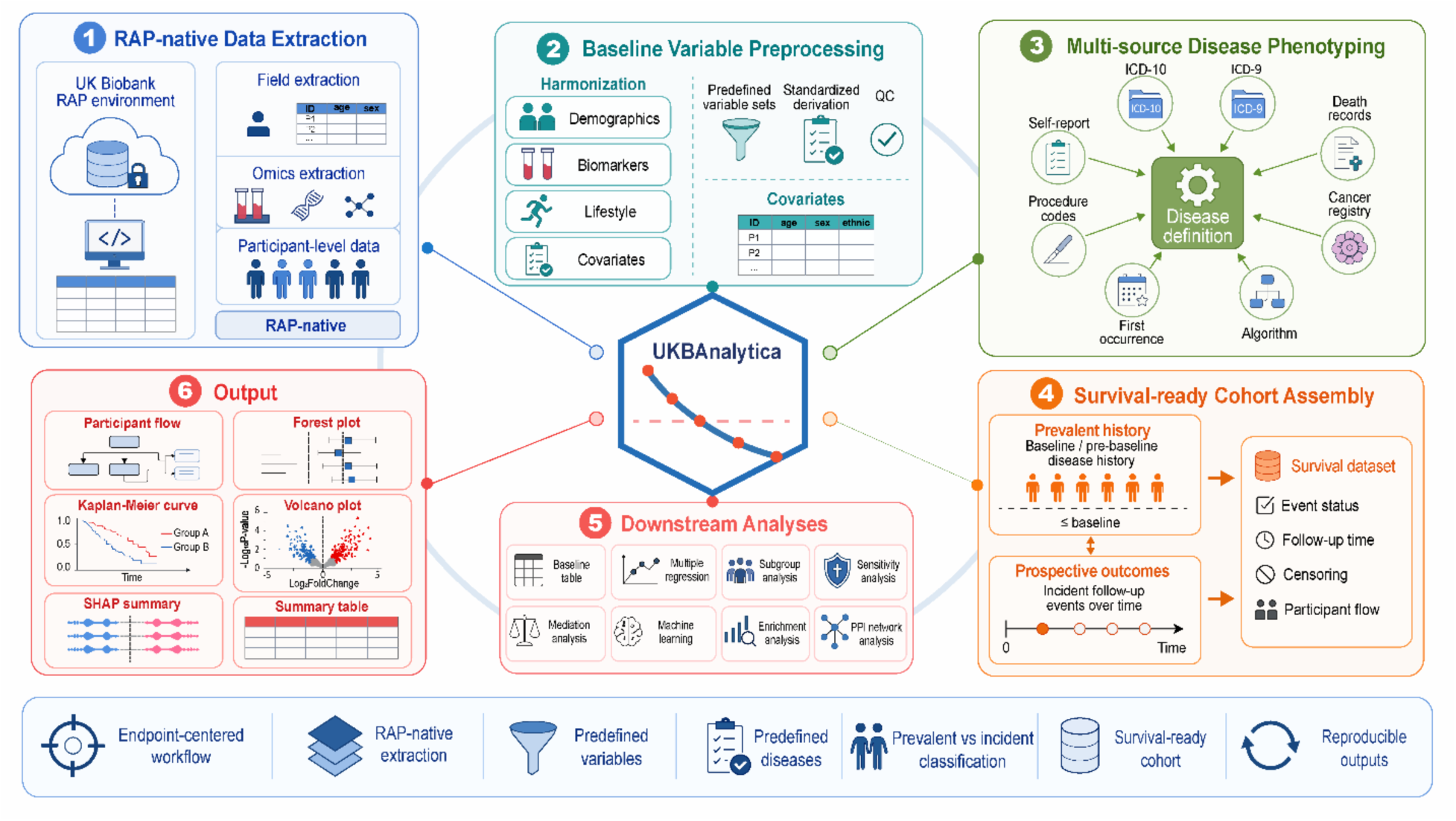
Overview of UKBAnalytica. The package provides six major functional modules, including RAP data extraction (online only), baseline data preprocessing, multi-source phenotyping definition, cohort assembly, downstream analyses, and visualization. **Notes:** The extraction module is designed for use within the RAP environment and does not require local download of individual-level raw data.

**Table 1.** Main functions in UKBAnalytica.

| Function | Description |
| --- | --- |
| rap_extract_pheno() | Extract phenotype fields or predefined variables from a UK Biobank RAP dataset using field IDs or package variable names. |
| get_variable_info() | Return predefined baseline variables that can be standardized by preprocess_baseline(). |
| preprocess_baseline() | Clean and harmonize predefined or user-defined baseline covariates from raw RAP-format fields. |
| get_variable_sets() | Return curated UK Biobank field sets for RAP extraction, including clinical, biomarker, environmental, family history, medication, and genetic fields. |
| get_predefined_diseases() | Retrieve curated multi-source disease definitions integrating different fields. |
| create_disease_definition() | Create a customized disease definition from diagnosis codes, self-report codes, procedure codes, cancer registry fields, or algorithmic outcome fields. |
| build_survival_dataset() | Construct survival-ready cohorts by classifying prevalent and incident cases, calculating follow-up time, and assigning censoring status. |
| run_regression() | Unified interface for linear, logistic, Cox, generalized linear, negative-binomial, and generalized additive models, with automatic dispatch to the corresponding runmulti_*() wrapper via the type argument. |
| ukb_ml_flow() | Run a complete single-model ML workflow. |
| ukb_ml_compare_flows() | Compare multiple feature sets, multiple ML algorithms, or their full combination using a unified ukb_ml_flow-based interface. |

The first layer covers remote data acquisition and standardized baseline preprocessing. Users can retrieve RAP phenotype fields through rap_extract_pheno() by specifying UK Biobank field IDs, predefined variable names, and their project-specific RAP dataset. The package currently includes 52 predefined baseline variables available through get_variable_info(), covering demographics, socioeconomic status, anthropometrics, blood pressure, lifestyle factors, medications, biomarkers, air pollution exposures, and diet-related variables. In addition, get_variable_sets() provides 15 curated field sets comprising 167 field-level entries across 143 unique UK Biobank field IDs, facilitating structured extraction of clinical core variables, biomarkers, environmental exposures, family history, self-reported diagnoses, medications, genetic principal components, and algorithmically defined outcomes. Practical preprocessing functions allow users to derive standardized covariates from raw RAP-format fields with minimal manual recoding.

The second layer centers on multi-source disease definition, prevalent-versus-incident classification, and survival-ready cohort construction. A key feature of this layer is its built-in library of 331 curated disease definitions, retrievable with get_predefined_diseases(). These definitions integrate multiple UK Biobank disease ascertainment sources, including ICD-10 and ICD-9 hospital diagnosis codes, self-reported illness codes, death registry ICD-10 codes, OPCS-4 operative procedure codes, UK Biobank First Occurrence fields, cancer registry records, and algorithmically defined outcome fields where available. The predefined endpoints span cardiovascular, metabolic, respiratory, renal, neurological, psychiatric, gastrointestinal, musculoskeletal, ophthalmological, dermatological, and cancer outcomes. For customized endpoints, create_disease_definition() and combine_disease_definitions() allow users to construct complex phenotype definitions dynamically. Subsequently, build_survival_dataset() processes these disease records to distinguish prevalent from incident cases, calculate follow-up time, and assign censoring status, thereby producing analysis-ready survival datasets.

The package also includes a flexible machine-learning framework for classification and survival modeling. Users can either run the complete modeling pipeline through a single high-level interface or execute each step separately for full analytical control. The ukb_ml_flow() function provides an end-to-end single-model workflow, covering data splitting, cross-validation, hyperparameter tuning, threshold selection, final model fitting, independent test-set evaluation, and optional SHAP-based interpretation. For comparative modeling, ukb_ml_compare_flows() extends this framework to compare different feature sets, different machine-learning algorithms, or their full combination under a unified interface. For users requiring stepwise implementation, lower-level functions such as ukb_ml_split_data(), ukb_ml_tune(), ukb_ml_threshold(), ukb_ml_fit_final(), ukb_ml_evaluate_test(), ukb_ml_roc_data(), and ukb_shap() remain available. Additional modules support causal mediation analysis, proteomic and metabolomic analyses, and publication-oriented visualizations.

### 2.2 Comparison with existing R packages related to UK Biobank data analysis

We next positioned UKBAnalytica relative to representative UK Biobank analysis packages, including ukbtools and PHESANT. These tools address important but distinct analytical needs (Table 2). ukbtools is an open-source R package designed mainly for local UK Biobank file management, data wrangling, ICD query, and genetic metadata handling. PHESANT is an open-source phenome-wide association scan pipeline that automates association testing across heterogeneous UK Biobank phenotypes. In contrast, UKBAnalytica was designed as a source-aware epidemiological workflow package that links RAP extraction, predefined variable preprocessing, multi-source disease definition, prevalent-versus-incident classification, survival-ready cohort construction, and downstream statistical or machine-learning analysis within a single R framework.

**Table 2.**
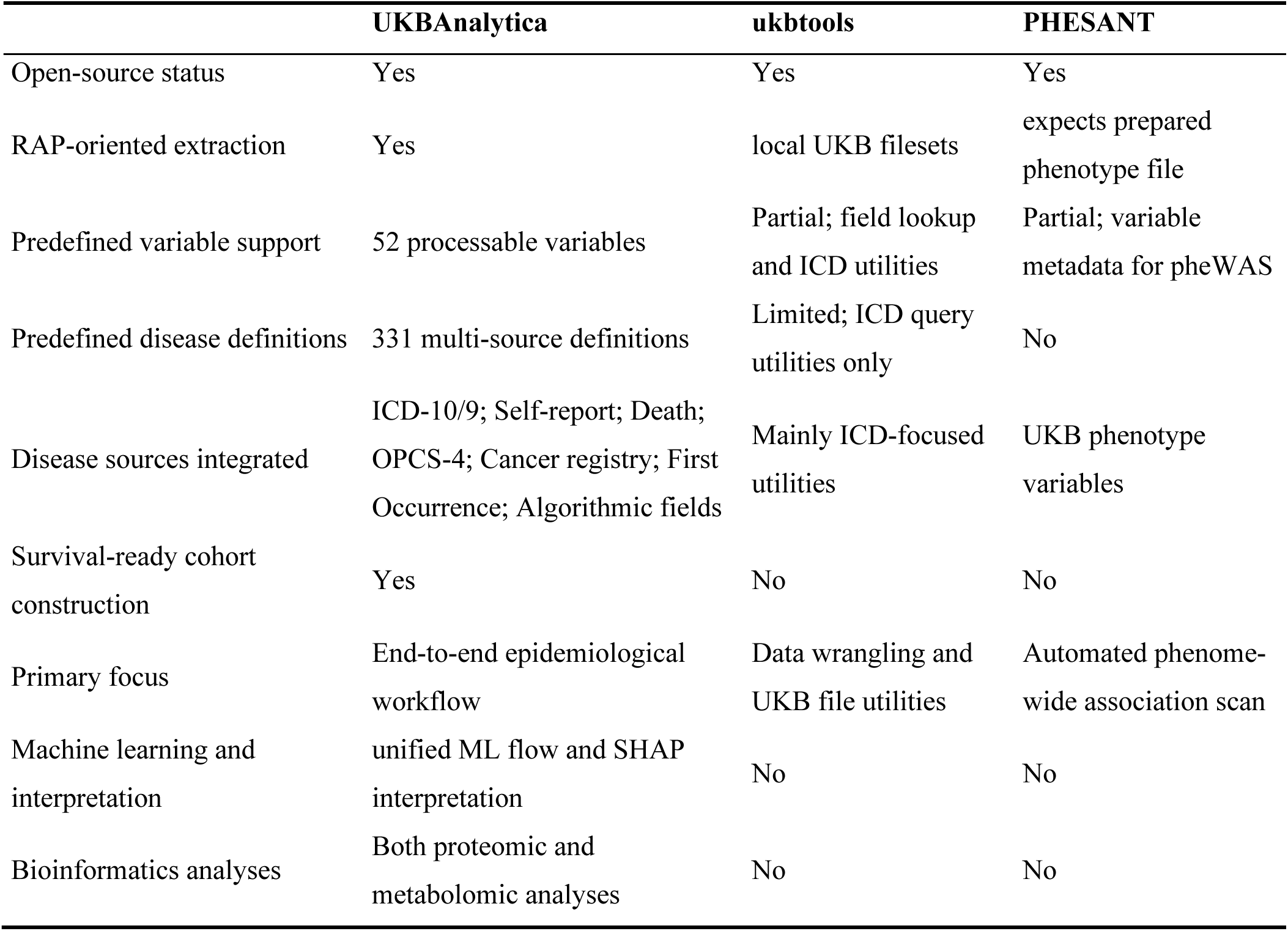
Feature comparison between UKBAnalytica and other representative UKB data analysis R packages.

### 2.3 Case study: end-to-end proteomic profiling of incident COPD using UKBAnalytica

#### 2.3.1 Cohort construction and phenotype harmonization

This case study was designed to demonstrate the practical use of UKBAnalytica in a RAP-based analysis workflow. Using COPD proteomic profiling as an example, we applied the package within the UK Biobank Research Analysis Platform and implemented an end-to-end pipeline covering data extraction, baseline variable preprocessing, phenotype definition, cohort construction, statistical modeling, machine-learning prediction, and downstream result visualization. This example illustrates how UKBAnalytica can standardize common UK Biobank analytical steps and connect phenotype curation with reproducible epidemiological and predictive modeling within a single R framework.

#### 2.3.2 Baseline characteristics of participants stratified by incident COPD status

A total of 50,172 participants were included in the main analytic cohort, including 2,427 incident COPD cases. The cohort was split into a training set of 35,121 participants and an internal validation set of 15,051 participants, with identical COPD event rates in both sets (4.84%).

Baseline characteristics differed significantly between participants with and without incident COPD (Table 3). Overall, participants who developed COPD were older, more likely to be male, had higher BMI and higher Townsend deprivation index values, and were more likely to be current or previous smokers (all P < 0.001). They also had lower educational attainment compared with participants who remained free of COPD. Similar baseline patterns were observed in the training and validation sets, supporting the comparability of the split (Table S1, Table S2).

**Table 3.** Baseline characteristics of participants stratified by incident COPD.

|  | Without COPD | With COPD | P |
| --- | --- | --- | --- |
| N | 47745 | 2427 |  |
| Age (mean (SD)) | 56.47 (8.22) | 61.10 (6.54) | <0.001 |
| Bmi (mean (SD)) | 27.38 (4.72) | 28.42 (5.50) | <0.001 |
| TDI (mean (SD)) | -1.30 (3.12) | -0.09 (3.54) | <0.001 |
| Sex (%) |  |  |  |
| Female | 26076 (54.6) | 1114 (45.9) | <0.001 |
| Male | 21669 (45.4) | 1313 (54.1) |  |
| Smoking status (%) |  |  |  |
| Never | 27093 (56.7) | 524 (21.6) | <0.001 |
| Previous | 16341 (34.2) | 1065 (43.9) |  |
| Current | 4311 (9.0) | 838 (34.5) |  |
| Ethnic (%) |  |  |  |
| White | 44745 (93.7) | 2350 (96.8) | <0.001 |
| Others | 3000 (6.3) | 77 (3.2) |  |
| Education (%) |  |  |  |
| Low | 7750 (16.2) | 938 (38.6) | <0.001 |
| Medium | 10625 (22.3) | 501 (20.6) |  |
| High | 29370 (61.5) | 988 (40.7) |  |

#### 2.3.3 Proteomic signatures associated with incident COPD

In the training set, Cox regression identified 163 proteins associated with incident COPD after Bonferroni adjustment among 2,920 tested proteins (Fig. 2a). The strongest positive associations included GDF15, CEACAM6, ALPP, CDCP1, SHISA5, AREG, CHCHD10, ADM, SPINT1, and FGF23, with GDF15 showing the strongest statistical signal (HR = 1.51, 95% CI: 1.46-1.56, adjusted P < 0.001). Several proteins showed inverse associations, including FAP, ADAMTS8, FASLG, CLEC3B, CEACAM16, FGFBP1, AGER, and ADAMTS13. In the validation set, 69 proteins remained significant after Bonferroni adjustment (Fig. 2b), with top positive and inverse associations shown in Fig. 2c and Fig. 2d. Among the 163 Bonferroni-significant proteins from the training set, 63 remained Bonferroni significant in validation, and 161 showed the same direction of association. Log hazard ratios were strongly correlated between training and validation for the training-set Bonferroni-significant proteins (Pearson r = 0.895; Spearman rho = 0.795), supporting the reproducibility of the COPD proteomic profile (Fig. S2).

**Fig. 2.**
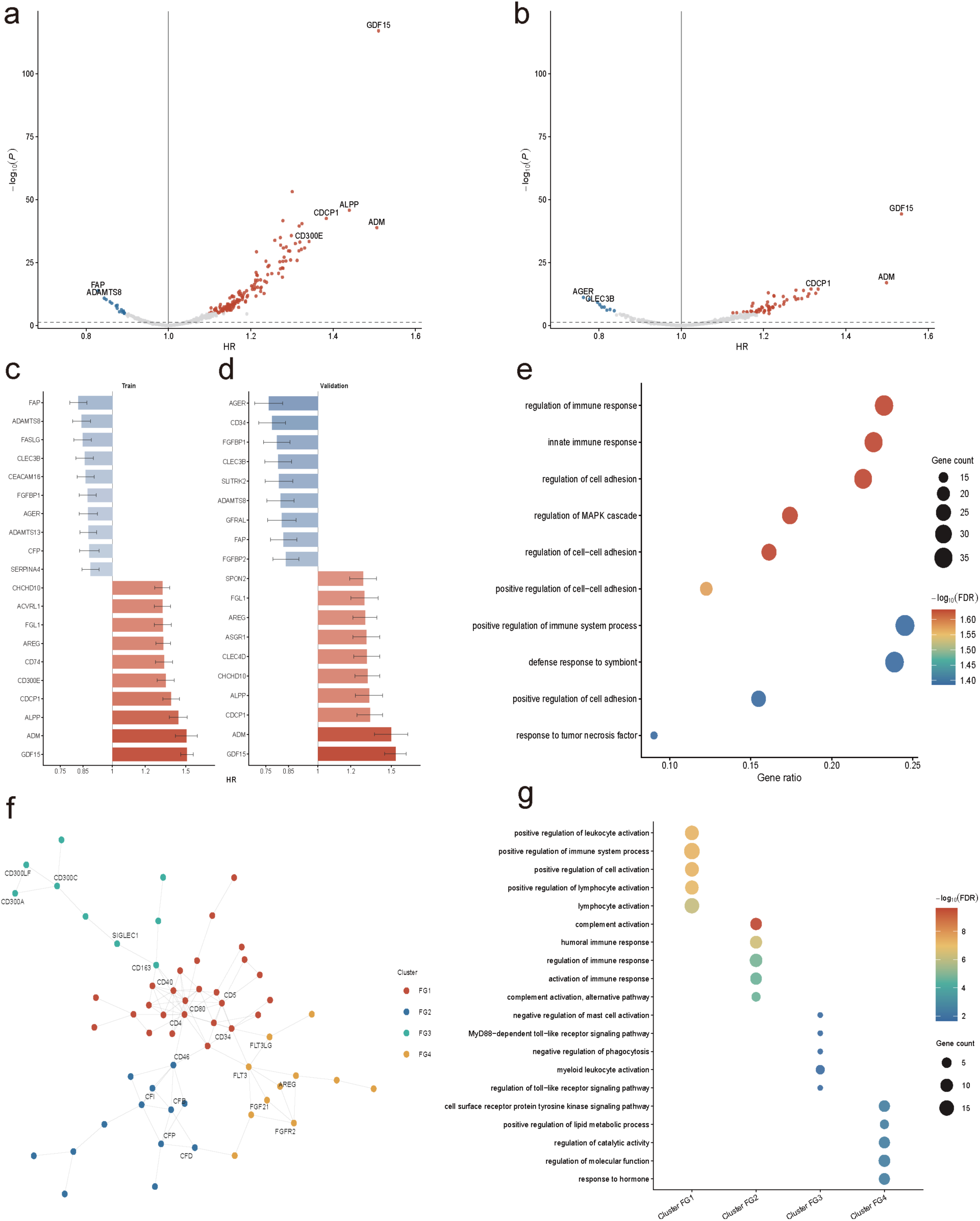
Proteomic profile of incident COPD. **a**, Volcano plot of protein-COPD associations in the training set. **b**, Volcano plot of protein-COPD associations in the validation set. **c**, Top positive and inverse COPD-associated proteins in the training set ranked by hazard ratio (HR). **d**, Top positive and inverse COPD-associated proteins in the validation set. **e**, GO biological process enrichment analysis of Bonferroni-significant proteins identified in the training set. **f**, STRING-based protein-protein interaction network clustered using fast greedy community detection. **g**, Cluster-specific GO biological process enrichment analysis.

GO-BP enrichment analysis of the 163 Bonferroni-significant proteins showed enrichment in immune and inflammatory processes, including innate immune response, regulation of immune response, regulation of cell adhesion, regulation of MAPK cascade, complement activation, and response to tumor necrosis factor (Fig. 2e). PPI analysis further identified a connected protein interaction network, and fast greedy community detection separated the network into four protein clusters (Fig. 2f, Fig. S3). Cluster-specific enrichment analysis suggested that these communities were related to distinct biological processes: FG1 was mainly related to leukocyte and lymphocyte activation, FG2 was enriched for complement activation and humoral immune response, FG3 was associated with myeloid leukocyte activation and toll-like receptor-related processes, and FG4 was enriched for receptor tyrosine kinase signaling, lipid metabolic regulation, and hormone response (Fig. 2g).

#### 2.3.4 Machine-learning prediction and model interpretation

XGBoost models using three feature sets were used to compare the predictive value of proteins, clinical covariates, and their combination. In the internal validation set, the protein-only model achieved an AUC of 0.804 (95% CI: 0.788-0.820), which was comparable to the covariate-only model (AUC = 0.800, 95% CI: 0.784-0.816; DeLong P = 0.588). The combined model of protein and clinical variables showed the best discrimination (AUC = 0.824, 95% CI: 0.808-0.839), with significantly higher AUC than both the protein-only model (DeLong P < 0.001) and the covariate-only model (DeLong P < 0.001) (Fig. 3a). At the Youden-index threshold, the combined model achieved a sensitivity of 0.753, specificity of 0.738, positive predictive value of 0.127, negative predictive value of 0.983, F1 score of 0.218, and Brier score of 0.150 (Table S3).

**Fig. 3.**
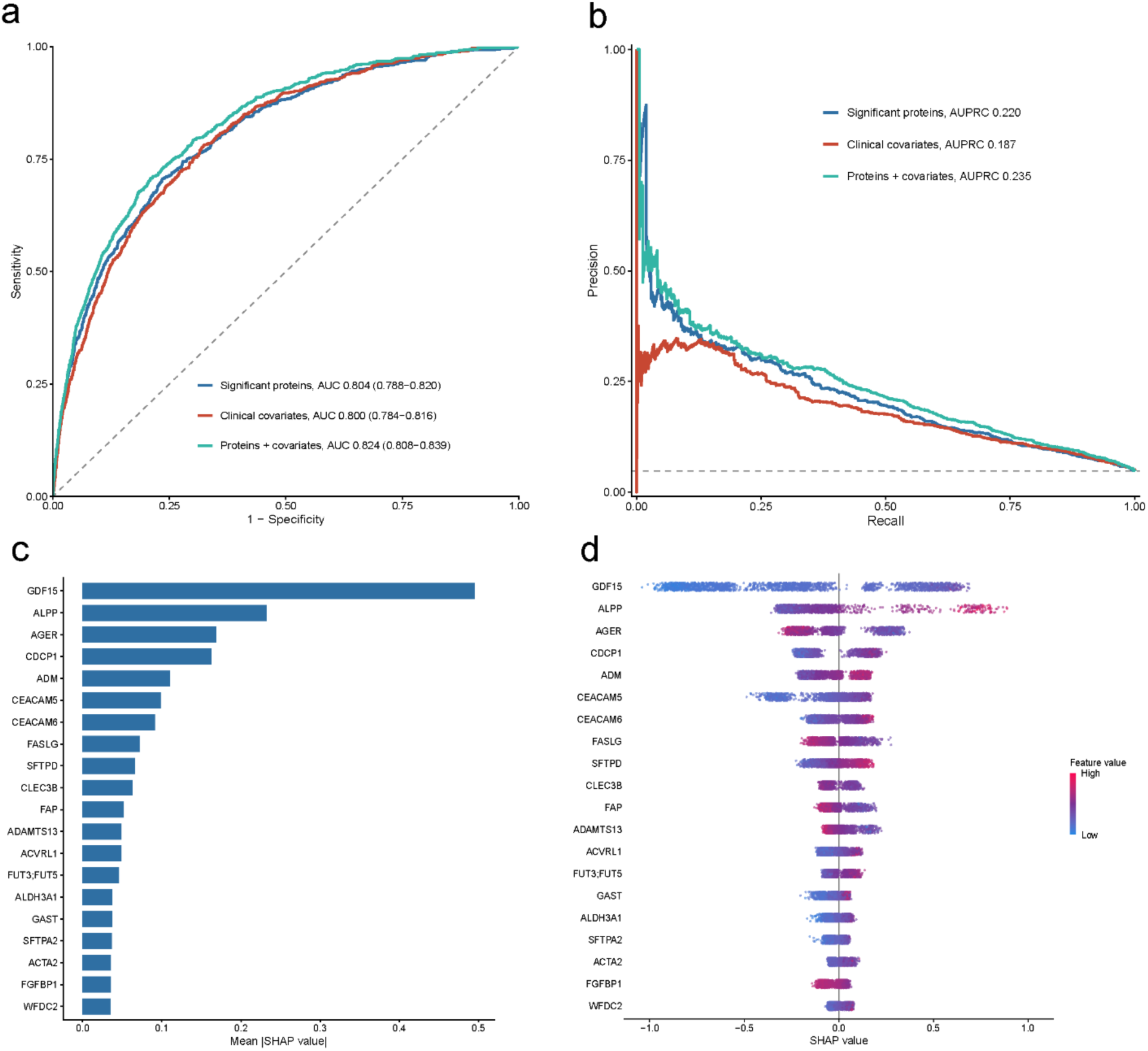
Machine-learning prediction and SHAP interpretation with COPD proteomics and clinical variables. **a**, Receiver operating characteristic curves comparing XGBoost models trained using proteins only, clinical covariates only, and proteins plus clinical covariates. **b**, Precision-recall curves for the three feature-set models. **c**, SHAP feature-importance bar plot for the protein-only model. **d**, SHAP beeswarm plot showing the magnitude and direction of protein contributions to COPD prediction.

Precision-recall analysis showed a similar pattern, with the highest AUPRC observed for the combined model (0.235), followed by the protein-only model (0.220) and the covariate-only model (0.187), all above the baseline event prevalence of 0.048 (Fig. 3b). Calibration and decision curve analyses are shown in Fig. S4. SHAP analysis of the protein-only model identified GDF15 as the most influential predictor, followed by ALPP, AGER, CDCP1, ADM, CEACAM5, CEACAM6, FASLG, SFTPD, CLEC3B, FAP, and ADAMTS13, indicating that the model prioritized proteins consistent with the Cox-based COPD proteomic profile (Fig. 3c, Fig. 3d).

#### 2.3.5 Risk stratification and lung-function relevance

Platt calibration was applied to the combined-model predictions using training-set out-of-fold predictions, and the calibrated probabilities were used for validation-set risk stratification. Participants were divided into low, intermediate, and high-risk groups according to calibrated predicted risk. Incident COPD rates increased stepwise across these groups, from 1.7% in the low-risk group to 7.0% in the intermediate-risk group and 22.7% in the high-risk group (Fig. 4a). Kaplan-Meier analysis showed clear separation of COPD-free survival across the three groups (Fig. 4b). Compared with the low-risk group, the intermediate- and high-risk groups had higher COPD hazards, with HRs of 4.52 (95% CI: 3.70-5.53) and 17.78 (95% CI: 14.82-21.34), respectively (Fig. 4c).

**Fig. 4.**
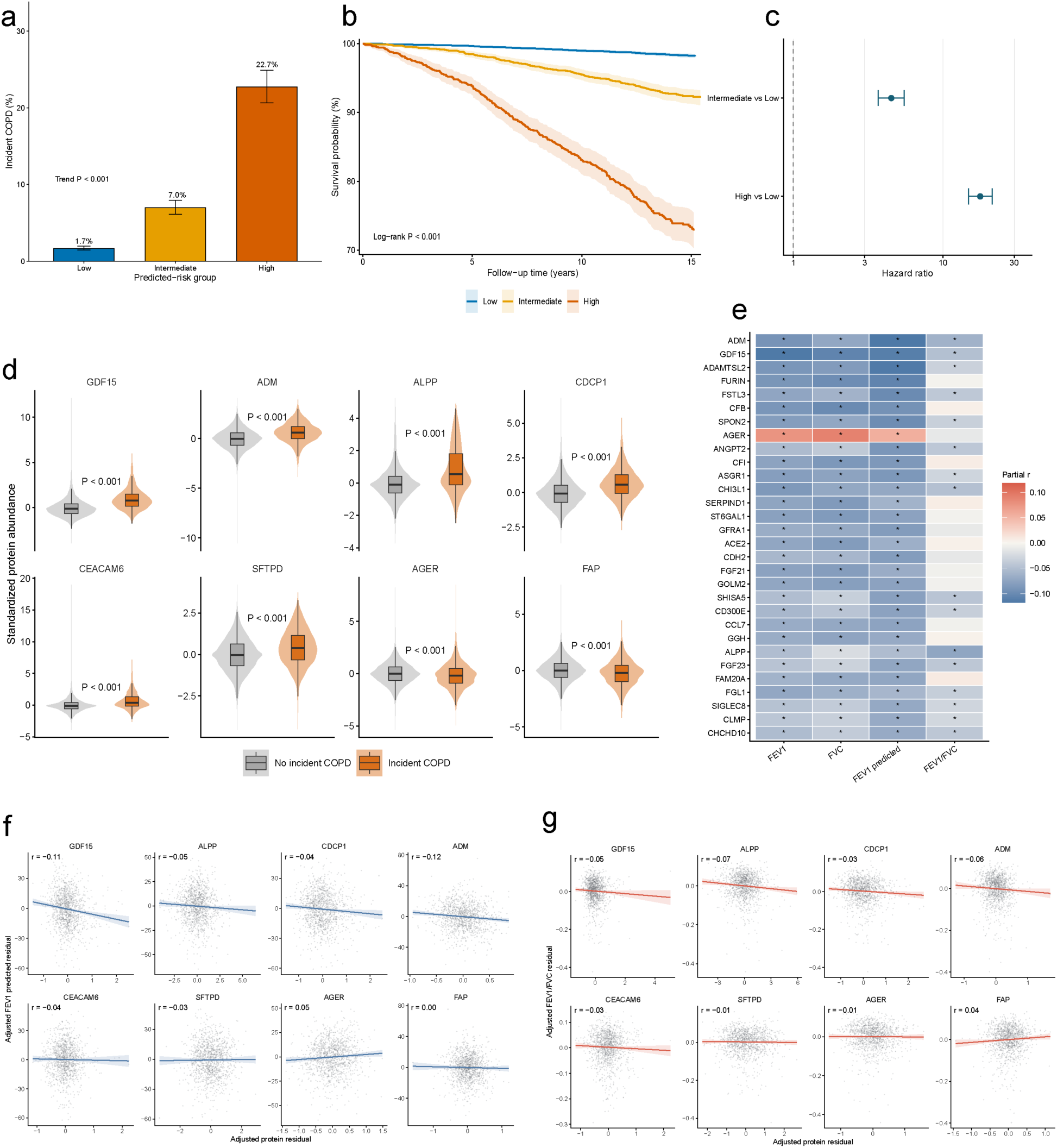
Risk stratification and lung-function relevance of COPD-associated proteins. **a**, Incident COPD proportions across calibrated predicted-risk groups in the validation set. **b**, Kaplan-Meier curves for COPD-free survival according to predicted-risk groups. **c**, Cox regression estimates comparing intermediate and high-risk groups with the low-risk group. **d**, Abundance distributions of representative COPD-associated proteins in participants with and without incident COPD. **e**, Partial correlation heatmap between Bonferroni-significant COPD proteins and spirometry-derived traits. **f**, Representative residualized scatter plots linking selected proteins with FEV1 percentage predicted. **g**, Representative residualized scatter plots linking selected proteins with FEV1/FVC ratio.

Representative COPD-associated proteins showed clear abundance differences between participants with and without incident COPD. Risk-increasing proteins, including GDF15, ADM, ALPP, CDCP1, CEACAM6, and SFTPD, were higher among incident COPD cases, whereas inverse-associated proteins such as AGER and FAP were lower among cases (Fig. 4d). To further relate the proteomic profile to lung-function phenotypes, partial correlation analyses were performed between the 163 Bonferroni-significant proteins and spirometry-derived traits after adjustment for baseline covariates. Many COPD-associated proteins showed modest but consistent correlations with FEV1, FVC, FEV1 percentage predicted, and FEV1/FVC ratio (Fig. 4e). Representative scatter plots showed that higher levels of risk-increasing proteins such as GDF15 and ADM were associated with lower FEV1 percentage predicted and FEV1/FVC, whereas AGER showed the opposite pattern for FEV1 percentage predicted (Fig. 4f, Fig. 4g).

#### 2.3.6 Sensitivity analyses

Sensitivity analyses supported the robustness of the protein-COPD associations. After excluding participants who developed COPD within the first 2, 4, and 6 years of follow-up, the log hazard ratios remained highly correlated with the main analysis (Pearson r = 0.991, 0.971, and 0.926, respectively), although the number of retained Bonferroni-significant proteins decreased as more early events were excluded (Fig. S5). Additional analyses showed similar consistency after adjustment for smoking pack-years (Pearson r = 0.955), adjustment for lung-function traits (Pearson r = 0.827), and exclusion of participants with baseline asthma (Pearson r = 0.958) (Fig. S6). Smoking-stratified analyses also showed positive correlations with the main analysis, with the strongest consistency among previous smokers (Pearson r = 0.879) (Fig. S6).

## 3. Discussion

In this study, we developed and evaluated UKBAnalytica as a phenotype-oriented R workflow for UK Biobank analyses conducted on the official Research Analysis Platform. The package was designed to start from endpoint and variable specification, rather than from isolated modeling functions, and to carry these definitions into reproducible cohort construction and downstream analysis. Using a COPD plasma proteomics case study on the RAP, we showed that this phenotype-oriented structure can support a complete applied workflow while maintaining a consistent analytical framework.

A key feature of UKBAnalytica is its structured handling of phenotypes and variables. The package currently provides 331 curated disease definitions and 52 predefined baseline variables, together with 15 curated variable sets comprising 167 field-level entries across 143 unique UK Biobank field IDs. These resources can be retrieved and inspected through get_predefined_diseases(), get_variable_info(), and get_variable_sets(), and then extended using create_disease_definition() and combine_disease_definitions() when a study requires customized endpoints. Together with preprocess_baseline() and build_survival_dataset(), this structure helps standardize covariate processing, prevalent and incident classification, follow-up time calculation, and censoring assignment. By making these definitions explicit and reusable, UKBAnalytica reduces repetitive recoding and lowers the risk of inconsistent endpoint construction across projects.

The downstream analysis modules are designed to connect naturally with the cohort outputs generated by the phenotype-construction workflow. After users define an endpoint and assemble an analysis-ready dataset, they can move within the same framework to epidemiological regression analysis, bioinformatics-oriented omics interpretation, and machine-learning prediction. Functions such as create_baseline_table(), runmulti_cox(), and ukb_ml_flow() allow these steps to be executed without repeatedly rebuilding model-specific input files. This smooth transition from phenotype definition to statistical modeling and reporting is central to the practical utility of UKBAnalytica.

UKBAnalytica should be considered complementary to existing UK Biobank tools rather than a replacement. Several existing tools address specific components of UK Biobank analysis. For example, ukbtools helps convert UK Biobank files into analysis-ready R objects, adds interpretable variable labels, supports ICD diagnosis queries, and handles genetic metadata^6^. PHESANT serves a different purpose by enabling automated phenome-wide association analyses across heterogeneous UK Biobank phenotypes^7^. ukbpheno focuses on semiautomated phenotyping of health-related outcomes by linking the main UK Biobank dataset with record-level health data files^9^. More recently, FastUKB has provided RAP-oriented functions for batch data extraction, graphical variable selection, automated cleaning, and baseline table generation, thereby reducing the technical burden of routine UK Biobank analyses^8^. Despite these strengths, existing tools still do not provide a complete workflow that connects data extraction, data preprocessing, cohort construction, and downstream modeling in a unified framework. This gap is particularly relevant for UK Biobank studies, as many commonly used analytical tools and statistical workflows are implemented in R^10–12^. In addition, many clinical and epidemiological machine-learning workflows are implemented in R^13,14^. Therefore, building an integrated R-based workflow ecosystem is important for improving reproducibility, reducing repeated manual coding, and supporting end-to-end analysis within a single environment. UKBAnalytica was developed to address this need. It links UK Biobank data extraction, baseline variable processing, endpoint definition, survival-ready cohort construction, and downstream epidemiological, omics, and machine-learning analyses. Its main contribution is to provide a transparent, scriptable, and phenotype-oriented R workflow for reproducible UK Biobank research.

The COPD proteomics case study served as a practical validation of whether UKBAnalytica could recover biologically credible signals from a RAP-based workflow. Our prioritized proteins were broadly consistent with previous COPD studies. For example, surfactant protein D has been reported as a circulating COPD biomarker and has been associated with smoking status and exacerbation risk^15,16^. Lower soluble RAGE, encoded by AGER, has been linked to COPD, reduced lung function, and greater emphysema severity, although its association with disease progression is less consistent^17^. CEACAM6 has been experimentally connected to cigarette smoke-related epithelial vulnerability and emphysema development, supporting its relevance to COPD susceptibility^18^. ADM-related biomarkers, especially MR-proADM or proADM, have also been associated with mortality and adverse outcomes in COPD^19,20^. The enriched pathways, including immune and inflammatory signaling, complement activation, MAPK-related pathways, and tissue remodeling, were also consistent with known COPD biology. Therefore, this case study suggests that UKBAnalytica can generate stable and biologically consistent results through a transparent and reproducible analytical pipeline.

Several limitations should be noted. First, although UKBAnalytica provides predefined variables and disease definitions, some highly specific or composite indicators still require study-specific operational definitions. A unified framework for all complex derived traits has not yet been implemented. Second, current phenotype definitions do not incorporate UK Biobank general practice (GP) records, which contain rich but heterogeneous primary-care information and require additional harmonization. Third, the package focuses on phenotype construction, epidemiological modeling, omics interpretation, and clinical prediction, but does not yet support large-scale genetic analyses such as GWAS or Mendelian randomization (MR). Future updates will extend disease and variable libraries, improve support for complex composite traits and additional health-record sources, and further connect UKBAnalytica with external tools to support a more complete end-to-end UK Biobank analysis workflow.

## 4. Conclusion

In summary, UKBAnalytica provides a phenotype-oriented, RAP-compatible R framework that links UK Biobank data extraction, variable preprocessing, multi-source endpoint definition, survival-ready cohort construction, and downstream analysis. It therefore offers a practical tool for researchers seeking transparent, reusable, and extensible UK Biobank analytical pipelines.

## 5. Methods

### 5.1 Overall framework and design objectives

UKBAnalytica was developed as a phenotype-centered R package for reproducible epidemiological analysis of UK Biobank data within the UK Biobank Research Analysis Platform (RAP). UK Biobank is a large population-based prospective cohort with extensive baseline assessment data and longitudinal linkage to multiple health-record sources, providing a major resource for investigating determinants of chronic diseases.

The package was designed to support three connected tasks that are often handled by separate project-specific scripts: the definition of phenotypes across heterogeneous health-record sources, the construction of survival-ready cohort datasets, and the execution of downstream analytical modeling. Rather than introducing new statistical estimators, UKBAnalytica provides a standardized workflow that maps RAP exports to reusable disease definitions, source-aware case ascertainment, and analysis-ready outputs. The implementation is modular, with dedicated components for data extraction, baseline preprocessing, disease definition, cohort assembly, downstream statistical analysis, and visualization (Fig. 1).

### 5.2 Multi-source phenotype definition framework

Each disease endpoint is represented as a reusable disease definition object that stores source-specific ascertainment rules. To build the predefined disease library, we manually curated disease identifiers and diagnostic codes from published literature and further incorporated publicly available phenotype definitions from PHESANT and an open-source Pomegranate library^7,21^. Totally, the current package provides 331 available disease diagnoses, which can be used directly or further customized by users. Supported evidence sources include hospital inpatient ICD-10 and ICD-9 diagnoses, self-reported illnesses, causes of death from death registry records, OPCS-4 procedure codes, records from the cancer registry, UK Biobank first-occurrence fields, and algorithmically defined outcomes when available. This abstraction allows the same disease to be represented either by predefined rule sets distributed with the package or by user-specified definitions built from regular expressions, code lists, and optional source or date fields. By separating disease knowledge from downstream analysis code, the framework facilitates transparent endpoint specification, reuse across studies, and the construction of composite outcomes from multiple component definitions.

### 5.3 Case extraction and survival dataset construction

For each disease definition, diagnosis evidence is first extracted separately from the user-selected sources and then harmonized to participant-level dates by retaining the earliest qualifying record for each participant. In the main cohort-construction workflow, disease ascertainment is explicitly divided into two layers. The first layer captures baseline and pre-baseline disease history and is controlled through the “prevalent_sources” argument. These records are used to generate disease-specific_history variables, which can be incorporated as covariates in downstream regression models to adjust for comorbidity burden at study entry. The second layer captures prospective outcomes during follow-up and is controlled through the “outcome_sources” argument. This design preserves the temporal ordering required for cohort analysis, because incident events are defined only from diagnoses occurring after the baseline assessment date.

Users can specify the source combination for each layer according to the scientific question and the expected precision of the underlying records, including ICD-10, ICD-9, self-report, death registry, OPCS-4 procedures, cancer registry records, first-occurrence fields, and algorithmically defined outcomes when available. In practice, self-reported conditions are often useful for identifying pre-existing disease at or before baseline, whereas prospective endpoint ascertainment generally relies more heavily on sources with explicit event dates. Participants are therefore classified as having prevalent disease when the earliest qualifying diagnosis date is on or before the baseline assessment date, and as having incident disease when the earliest qualifying diagnosis date from the outcome layer occurs after baseline. Participants with prevalent disease were excluded from the risk set for the corresponding incident endpoint, even if subsequent records were present during follow-up. For the primary endpoint, individuals with prevalent disease are removed from the at-risk set by assigning missing values to “outcome_status” and “outcome_surv_time”, whereas non-prevalent individuals are followed until the earliest of the event date, the date of death, or the administrative censoring date. Follow-up time is then calculated as t = (d − b)/365.25 for incident cases and t = (min(c, m) − b)/365.25 for censored participants, where b denotes the baseline date, d denotes the diagnosis date, c denotes the administrative censor date, and m denotes the date of death. The final output retains the full cohort and appends disease-specific history and incident indicators together with standardized outcome variables for downstream survival modeling.

### 5.4 Downstream analytical modules

The downstream analysis layer was organized into three major components: epidemiological analysis, omics-oriented analysis, and machine-learning-based modeling and interpretation. Additional utilities support multiple imputation, propensity score analysis, mediation analysis, and publication-oriented visualization.

#### 5.4.1 Baseline descriptive summaries

Baseline descriptive summaries are generated with create_baseline_table(), which prepares the case indicator and selected covariates by coercing grouping and categorical variables to factor format and continuous variables to numeric format before calling CreateTableOne() from the tableone package. This wrapper standardizes the creation of baseline characteristic tables from case-control or exposed-unexposed comparisons and allows optional hypothesis testing within the same workflow.

#### 5.4.2 Regression modeling and diagnostics

Association analyses are supported through the unified run_regression() interface, which dispatches to six model-specific wrappers according to the type argument, including runmulti_cox(), runmulti_logit(), runmulti_lm(), runmulti_glm(), runmulti_negbin(), and runmulti_gam(). These functions iterate over user-specified exposures, construct model formulas automatically, and return harmonized tables of effect estimates, 95% confidence intervals, and P values. Supported outputs include hazard ratios, odds ratios, regression coefficients, and incidence rate ratios, depending on the model type.

Generalized linear models support common model families such as Poisson, Gamma, and quasi-likelihood models; negative-binomial models are used for overdispersed count outcomes; and generalized additive models fit penalized spline terms using gam() from the mgcv package to assess potential non-linear exposure-outcome relationships^22^. For time-to-event analyses, runmulti_cox_zph() and runmulti_trend() provide proportional-hazards diagnostics and grouped trend testing within the same analytical interface.

#### 5.4.3 Subgroup and interaction analysis

Subgroup analyses are implemented through run_subgroup_analysis() and run_multi_subgroup(). These functions first evaluate the exposure-by-subgroup interaction in the full model, using likelihood-ratio tests for multi-level subgroups and Wald tests for binary subgroups, and then refit the selected model within each subgroup level. Supported model types include Cox, logistic, linear, generalized linear, and negative-binomial models. The outputs include subgroup-specific sample size, event count, effect estimate, 95% confidence interval, and interaction P value, and can be directly visualized using plot_forest().

#### 5.4.4 Sensitivity-analysis preprocessing

Sensitivity analyses are implemented as preprocessing modules that preserve compatibility with the main regression wrappers. sensitivity_exclude_early_events() removes participants who experience the endpoint within a user-specified lag period after baseline, whereas sensitivity_exclude_missing_covariates() excludes observations with incomplete adjustment variables. Both functions preserve the original columns and object class of the analysis dataset and attach sensitivity_info metadata for auditability. In addition, runmulti_cox_lag() combines early-event exclusion with repeated Cox refitting to support lagged survival analyses.

#### 5.4.5 Multiple imputation and pooled inference

Multiple imputation is initiated through run_imputation(), which wraps mice() from mice package to impute a selected set of covariates while leaving identifier, exposure, outcome, and follow-up variables unchanged^23^. The function returns both the object and a list of completed datasets merged back to the static cohort skeleton, which is useful when only selected covariates require imputation. Downstream pooling is handled by fit_mi_models(), create_imputation_list(), and pool_mi_models(), which fit linear, logistic, Poisson, Cox, or negative binomial models through stats, survival, and MASS backends and combine estimates with MIcombine() from mitools package according to Rubin’s rules^24^.

#### 5.4.6 Propensity score analysis

Propensity score workflows are implemented through estimate_propensity_score(), match_propensity(), calculate_weights(), assess_balance(), and run_weighted_analysis(). Propensity scores can be estimated either by logistic regression using glm() or by gradient boosting using the gbm package; matched cohorts are generated through matchit() from MatchIt; inverse-probability weights for ATE, ATT, or ATC estimands are computed directly within the package, with options for stabilization and truncation. Covariate balance is quantified with standardized mean differences and variance ratios, and weighted outcome models are fitted with coxph(), glm(), or lm(), with optional robust variance estimation from the sandwich and lmtest packages.

#### 5.4.7 Mediation analysis

Causal mediation analysis is provided by run_mediation(), which wraps regmedint() and supports continuous or binary mediators together with linear, logistic, or Cox outcome models^25^. The wrapper standardizes input validation, specification of exposure contrasts and covariate reference values, and extraction of controlled direct, natural direct, natural indirect, total, and proportion-mediated effects into a common result object. For higher-throughput analyses, run_multi_mediator() applies the same workflow iteratively across a user-defined mediator panel, and associated visualization functions summarize both single-mediator and multi-mediator decompositions.

#### 5.4.8 Machine learning and model interpretation module

Machine learning for binary, multiclass, and continuous outcomes is implemented through ukb_ml_flow() and ukb_ml_workflow(). These workflows cover data splitting, feature selection, model tuning, test-set evaluation, and SHAP-based interpretation. Supported learners include logistic and linear regression, random forests, xgboost, glmnet, support-vector machines, neural networks, decision trees, and naive Bayes. Users can inspect available algorithms with ukb_ml_supported_models(), compare feature sets or models with ukb_ml_compare_feature_sets() and ukb_ml_compare_flows(), and visualize performance using ROC, PR, calibration, DCA, and SHAP plotting helpers.

For time-to-event prediction, ukb_ml_survival_workflow() provides a parallel workflow for survival ML. It supports Cox models, regularized Cox models, random survival forests, and gradient-boosting survival models, with frozen-test evaluation based on Harrell C-index and time-specific prediction summaries. Stepwise functions for splitting, tuning, final fitting, evaluation, and survival SHAP interpretation are also available for users who need more granular control.

Model interpretability is supported through SHAP-based analysis. The ukb_shap() function computes SHAP values for fitted UKBAnalytica machine-learning workflow objects, while ukb_shap_summary(), plot_shap_beeswarm(), plot_shap_dependence(), and plot_shap_force() provide standardized summaries and visualizations of global and feature-specific model contributions^26^.

#### 5.4.9 Visualization and reporting outputs

Publication-oriented graphics are generated through a common visualization layer built primarily on ggplot2. The package supports forest plots, Kaplan-Meier curves, regression volcano plots, propensity-score diagnostics, mediation summaries, multiple-imputation diagnostics, and enrichment visualizations. For machine-learning workflows, the visualization module further provides ROC and precision-recall curves, calibration plots, decision-curve analysis, model-comparison plots, and SHAP-based beeswarm, summary, dependence, and force plots. These functions allow outputs from the core statistical and machine-learning wrappers to be translated directly into manuscript-ready figures with minimal additional reshaping. Multi-panel figure assembly is supported through the aplot package^27^.

#### 5.4.10 Skills for online UK Biobank data analysis

To support AI-assisted coding and result organization, UKBAnalytica provides a provider-agnostic skill pack that converts major package functions into reusable agent skills. The skill pack includes 14 skills covering RAP data extraction, phenotype construction, preprocessing, epidemiological analysis, machine learning, proteomics, visualization, and workflow orchestration. Each skill contains a concise description, function references, and executable examples to help researchers write analysis code and summarize outputs more efficiently. Importantly, these skills are designed as workflow aids rather than data-access tools. The skill instructions explicitly restrict AI agents from interacting with participant-level RAP data or direct identifiers. Individual-level data must remain within the approved RAP project, whereas only aggregate results, code templates, and de-identified figures may be used for reporting. This design reduces the risk of data leakage while preserving the convenience of agent-assisted analysis.

### 5.5 Case study data preprocessing

#### 5.5.1 Study Population

This study used data from the UK Biobank, a large-scale prospective cohort that recruited more than 500,000 participants from 22 assessment centers across England, Scotland, and Wales between 2006 and 2010. Baseline information on socioeconomic characteristics, lifestyle factors, and disease history was collected for all participants. The UK Biobank study received ethical approval from the North West Multi-center Research Ethics Committee (approval number: 11/NW/0382), and all participants provided written informed consent.

Participants were excluded if they had missing baseline covariate information, unavailable plasma proteomic data, prevalent COPD at baseline, or incomplete outcome information. We also excluded individuals with invalid follow-up time, including those with missing or non-positive follow-up duration. After these exclusions, the remaining participants were included in the final analytical cohort for subsequent proteomic and downstream survival analyses (Figure S1).

#### 5.5.2 Proteomics assays

Plasma samples collected at baseline were stored by UK Biobank under standardized biobank conditions, including −80 °C storage and liquid nitrogen preservation. Proteomic profiling was performed using the Olink proximity extension assay platform, which enables high-throughput quantification of circulating plasma proteins. Protein measurements were generated across multiple Olink panels, including cardiometabolic, inflammation, neurology, and oncology-related proteins. Detailed information regarding assay design, analytical performance, and validation procedures has been described by UK Biobank and Olink elsewhere^28^.

Stringent quality control procedures were applied before downstream analyses. Proteins with more than 50% missing measurements were excluded to reduce the influence of poorly detected analytes. After this filtering step, 2,920 plasma proteins were retained for subsequent analyses. For the remaining proteins with incomplete measurements, missing values were imputed using the k-nearest neighbors (KNN) algorithm with k = 10. The final imputed proteomic dataset was then used for downstream regression, feature selection, and predictive modeling analyses.

#### 5.5.3 Definition of COPD and survival time calculation

Chronic obstructive pulmonary disease (COPD) was defined using multiple UK Biobank data sources. The diagnostic criteria were based on ICD-10 codes J40–J44 and ICD-9 codes 491, 492, 493.2, and 496. Self-reported COPD-related conditions were identified using UK Biobank self-report codes 1112, 1113, and 1472. Algorithmically defined COPD records and first-occurrence fields were also incorporated to improve case ascertainment.

For this COPD case study, source selection was defined separately for baseline disease history and prospective outcome ascertainment. Baseline prevalent COPD was identified using ICD-10 records, self-reported diagnoses, and algorithmically defined outcomes to maximize detection of pre-existing disease before or at baseline. Prospective incident COPD was identified using ICD-10 and ICD-9 hospital records, death registry records, and algorithmically defined outcomes, which provide dated clinical events suitable for time-to-event analysis. Self-reported COPD was not used for prospective outcome ascertainment owing to limited event-date precision. Follow-up time was constructed using the build_survival_dataset() function in the UKBAnalytica R package. Participants were followed from baseline assessment until the first COPD event, death, or administrative censoring on October 7, 2025, whichever occurred first.

#### 5.5.4 Covariates

All covariates included in this study can be calculated using preprocess_baseline() automatically in the UKBAnalytica. The following covariates were included: age, sex, body mass index (BMI), smoking status, Townsend deprivation index (TDI), ethnicity, and education level. Age was treated as a continuous variable. Sex was categorized as male or female. BMI was calculated as weight divided by height squared and analyzed as a continuous variable. Smoking status was categorized as never, previous, or current smoker. TDI was included as a continuous indicator of area-level socioeconomic deprivation, with higher values indicating greater deprivation. Ethnicity was categorized as white and other ethnic groups, while education levels were categorized as low (none of the listed qualifications), medium (school-level qualifications), and high (college/university, vocational, or professional qualifications), with the highest attained category assigned when multiple qualifications were reported. All covariates were measured at baseline and included in multivariable models to reduce potential confounding.

#### 5.5.5 Statistical analysis

Baseline characteristics were summarized according to incident COPD status. Continuous variables were presented as mean with standard deviation or median with interquartile range, and categorical variables were summarized as counts and percentages. Between-group comparisons were performed using analysis of variance or Kruskal-Wallis tests for continuous variables and chi-square tests for categorical variables. Baseline tables were generated using the create_baseline_table() function in UKBAnalytica, which wraps the tableone R package. Participants with missing values in the primary adjustment covariates were excluded using a complete-case approach implemented by sensitivity_exclude_missing_covariates(stepwise = TRUE), and the stepwise exclusion flow was recorded. The final analytic cohort was randomly divided into the training and internal validation sets at a 7:3 ratio using ukb_ml_split_data().

Protein-COPD associations were first evaluated in the training set using Cox proportional hazards regression models implemented by runmulti_cox() in UKBAnalytica, which calls coxph() function from survival package. Each protein was standardized using the training-set mean and standard deviation, and the same scaling parameters were applied to the validation set. Models were adjusted for age, sex, body mass index (BMI), smoking status, Townsend deprivation index (TDI), ethnicity, and education. Hazard ratios, 95% confidence intervals, and P values were estimated for each protein, followed by multiple-testing correction using Bonferroni correction. Proteins significant in the training set were evaluated in the internal validation set, and concordance was assessed by replication proportions and Spearman correlations of log hazard ratios between training and validation results. Gene Ontology biological process enrichment analysis was performed for Bonferroni-significant proteins using clusterProfiler^29^. Protein-protein interaction networks were retrieved from STRING using get_protein_ppi(), topological metrics were computed using UKBAnalytica, and fast greedy community detection was performed using run_protein_ppi_fastgreedy(). Cluster-specific biological processes were further evaluated using compareCluster() from clusterProfiler^30^, and network and enrichment results were visualized with ggraph, ggplot2, and ComplexHeatmap.

XGBoost models were constructed to predict incident COPD using the machine-learning workflow implemented in UKBAnalytica. Three feature sets were evaluated: Bonferroni-significant proteins only, clinical covariates only, and Bonferroni-significant proteins plus clinical covariates. Model development was performed exclusively in the training set. Hyperparameters were optimized using 10-fold cross-validation with grid search by function ukb_ml_tune_grid(), and the final model was refitted on the full training set using the selected parameter combination. The internal validation set was held out during model tuning and was used only for independent model performance assessment with ukb_ml_evaluate_test(). The optimal classification threshold was selected from the training-set cross-validation predictions using the Youden index through ukb_ml_threshold() and then applied unchanged to the validation set. Model performance was evaluated using the area under the receiver operating characteristic curve (AUC), accuracy, sensitivity, specificity, positive predictive value, negative predictive value, F1 score, and Brier score (all metrics can be generated by ukb_ml_metrics()). Precision-recall curves, calibration plots, and decision curve analysis were additionally generated to evaluate performance under outcome imbalance, agreement between predicted and observed risk, and clinical net benefit. AUCs between models were compared using DeLong tests in the internal validation set. Platt scaling was fitted using training-set out-of-fold predictions and then applied to the validation-set predictions for calibrated risk estimation.

Model interpretability was assessed using SHAP values. SHAP values were calculated for the final XGBoost models to quantify the contribution of each feature to individual-level predictions. Feature importance was summarized using mean absolute SHAP values, and SHAP beeswarm plots from plot_shap_beeswarm() were used to visualize both the magnitude and direction of feature effects. For the combined model, calibrated predicted probabilities were used to stratify validation-set participants into low, intermediate, and high-risk groups. Incident COPD proportions, Kaplan-Meier survival curves, and Cox proportional hazards models after adjusting covariates were used to evaluate risk separation across predicted-risk groups.

To explore the relationship between COPD-associated proteins and baseline lung function, we performed partial correlation analyses between the 163 Bonferroni-significant COPD proteins and spirometry-derived traits, including FEV1, FVC, FEV1 percentage predicted, and FEV1/FVC ratio. For each protein-trait pair, both the protein abundance and lung-function trait were residualized against age, sex, BMI, smoking status, Townsend deprivation index, ethnicity, and education, and Pearson correlations were then calculated between the residuals. P values were adjusted using the Benjamini-Hochberg method.

Sensitivity analyses were performed to examine the robustness of the relationship between proteins and COPD. First, to reduce potential reverse causation, participants who developed COPD within the first 2, 4, or 6 years of follow-up were sequentially excluded, and protein-wide Cox regression analyses were repeated in the training set. The resulting log hazard ratios were compared with the main analysis using Pearson and Spearman correlations, and the proportion of Bonferroni-significant proteins retained after each exclusion was calculated. Additional sensitivity analyses assessed potential residual confounding by smoking burden and clinical heterogeneity, including additional adjustment for smoking pack-years, exclusion of participants with baseline asthma, and subgroup analyses by smoking status and sex where event counts were sufficient.

## Data availability

The data that support the findings of this study are available from UK Biobank (https://www.ukbiobank.ac.uk/). Individual-level UK Biobank data are available to approved researchers through the UK Biobank Research Analysis Platform under standard access procedures. The COPD proteomics case study was conducted under UK Biobank application number 822932. No participant-level data are distributed with UKBAnalytica.

## Code availability

The source code of UKBAnalytica is available at https://github.com/Hinna0818/UKBAnalytica. Full documentation, including function references and tutorials, is available at https://hinna0818.github.io/UKBAnalytica/.

## Acknowledgements

This research was conducted using data from the UK Biobank Resource under Application Number 822932. The authors gratefully acknowledge the UK Biobank participants and coordinators for their valuable contributions.

## Ethics Approval

All procedures were conducted in accordance with the Declaration of Helsinki. The UK Biobank study received ethical approval from the North West Multi-Centre Research Ethics Committee (REC reference: 11/NW/0382) and obtained informed consent from all participants. Access to the UK Biobank resource was granted under application number 822932 for the present analysis.

## Author contributions

N.H. and F.H. conceptualized the project. N.H. and G.Y. developed the UKBAnalytica package and performed the main analyses. G.Y. and K.M. reviewed and revised the code. N.H. and F.H. drafted the manuscript. All authors reviewed and approved the final manuscript.

## Competing interests

The authors declare no competing interests.

---

**Fig. S1.**
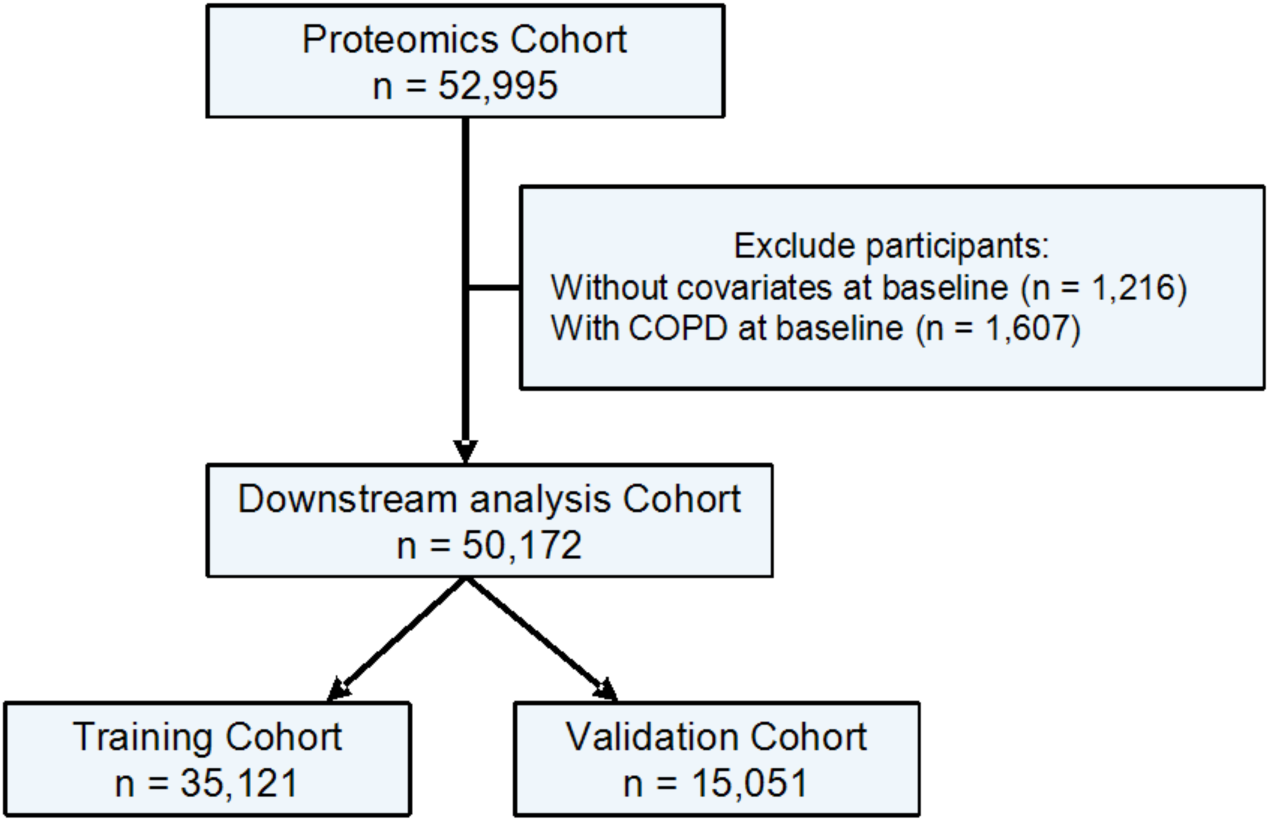
Participant flow for the COPD proteomics case study. Participants without baseline covariates records, or with COPD were excluded out of the downstream analyses. A total of 35,121 participants were kept in the training cohort, while 15,051 in the validation cohort.

**Fig. S2.**
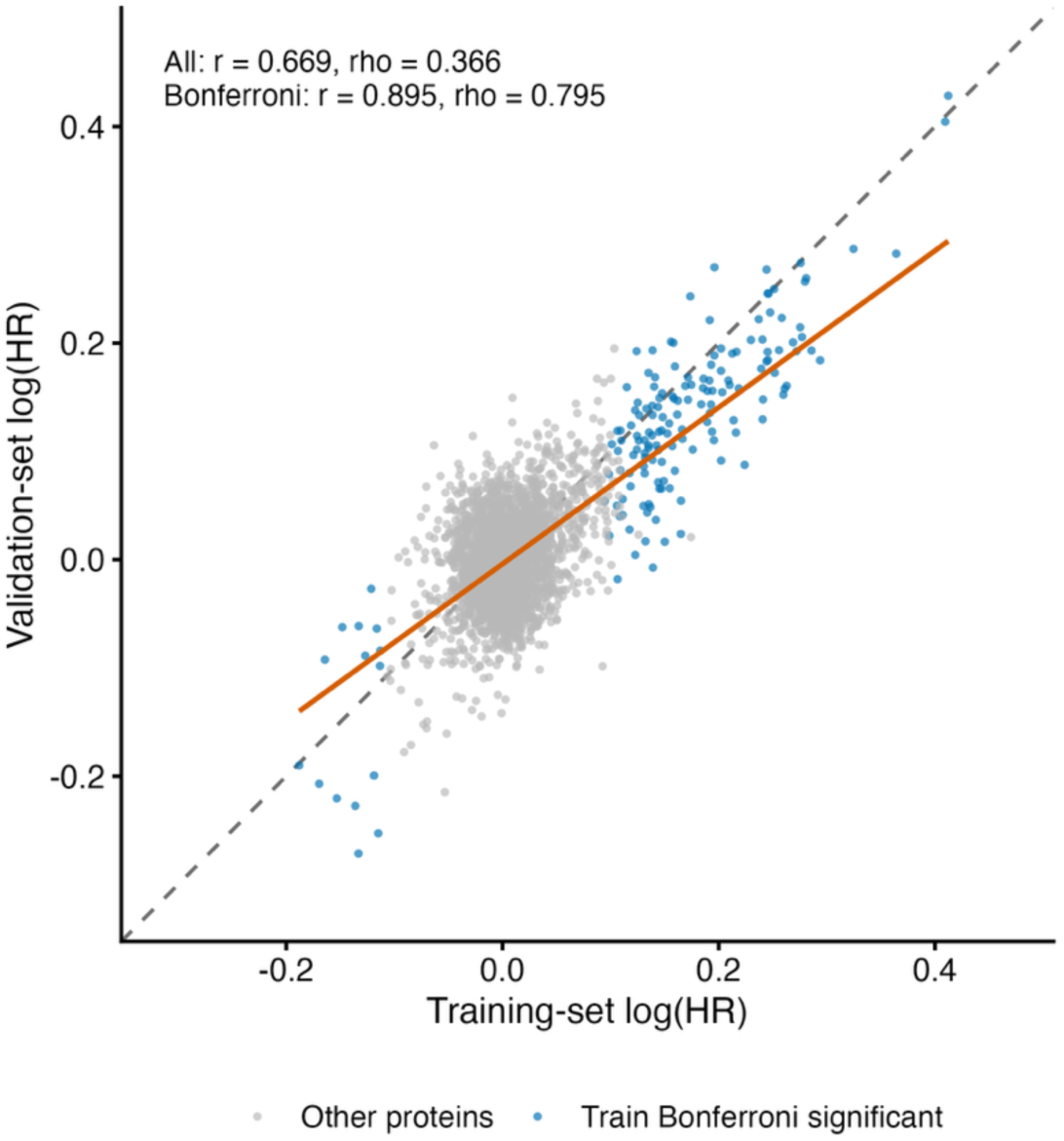
Correlation of protein-COPD associations between the training and validation sets. Each point represents one protein tested in the Cox regression analysis. The x axis shows the log hazard ratio (HR) estimated in the training set, and the y axis shows the corresponding log (HR) estimated in the validation set. Proteins significant after Bonferroni correction in the training set are highlighted. Pearson and Spearman correlation coefficients were calculated to evaluate the concordance of effect estimates across datasets.

**Fig. S3.**
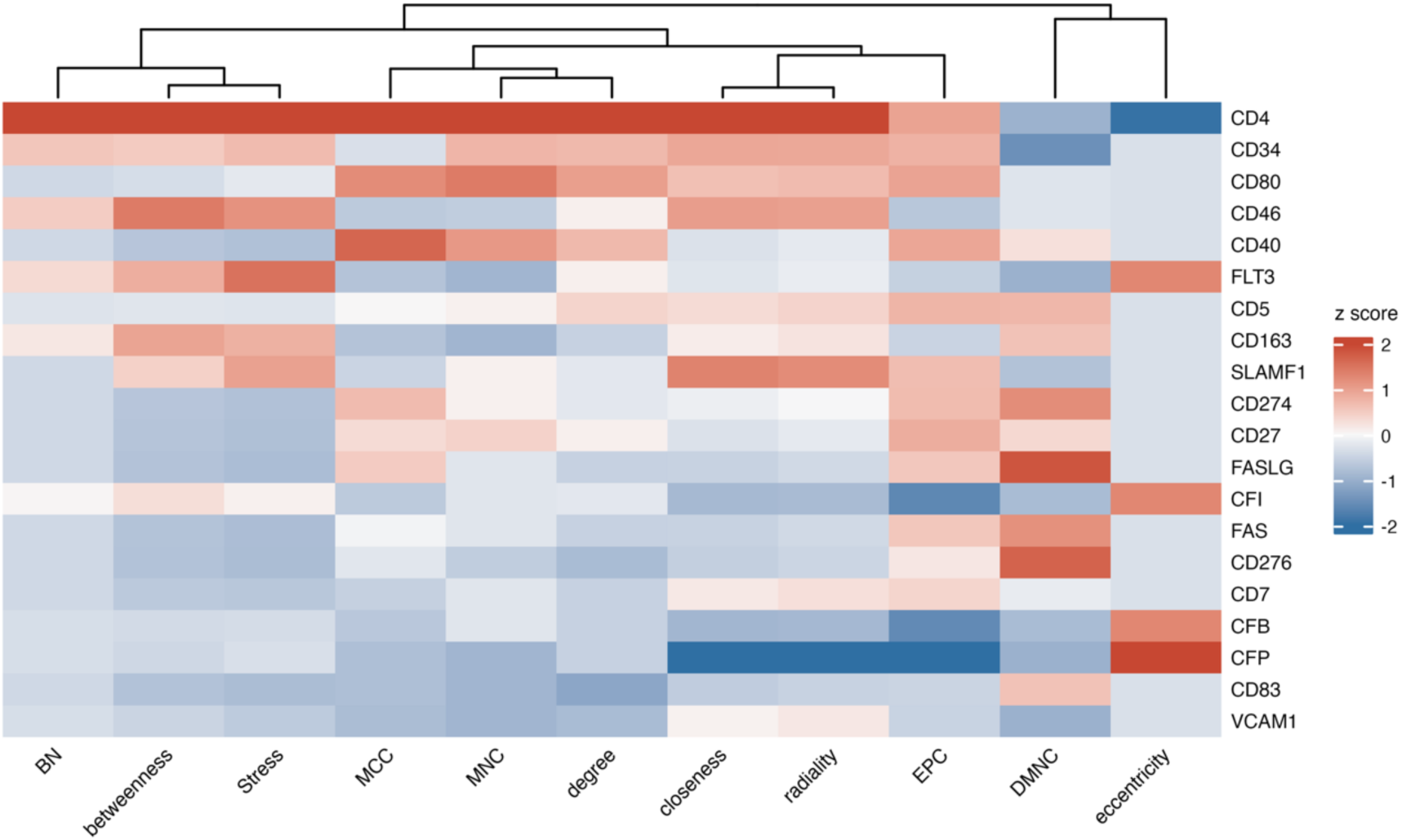
Standardized topological profiles of the top 20 comprehensively ranked protein nodes. The top 20 protein nodes were selected according to the integrated ranking score derived from protein-protein interaction network topology, and all topological metrics were standardized to enable cross-metric comparison.

**Fig. S4.**
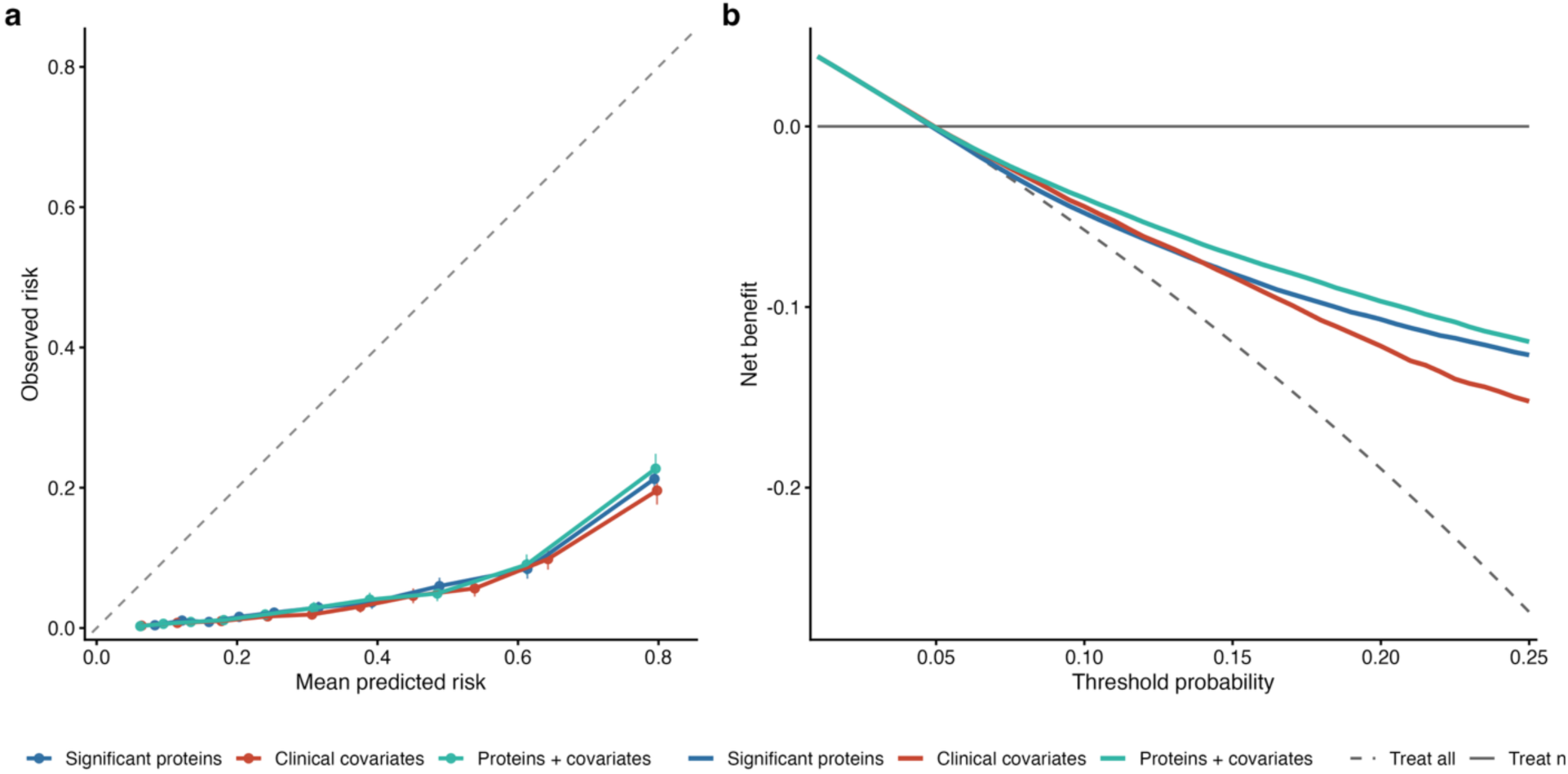
Calibration and decision curve analyses of XGBoost models in the validation set. **a**, Calibration curves compare mean predicted risks with observed incident COPD proportions across risk deciles for models trained using significant proteins only, clinical covariates only, and proteins plus clinical covariates. The dashed diagonal line indicates perfect calibration. **b**, Decision curve analysis shows the net benefit of the three models across clinically relevant threshold probabilities, with treat-all and treat-none strategies shown as references.

**Fig. S5.**
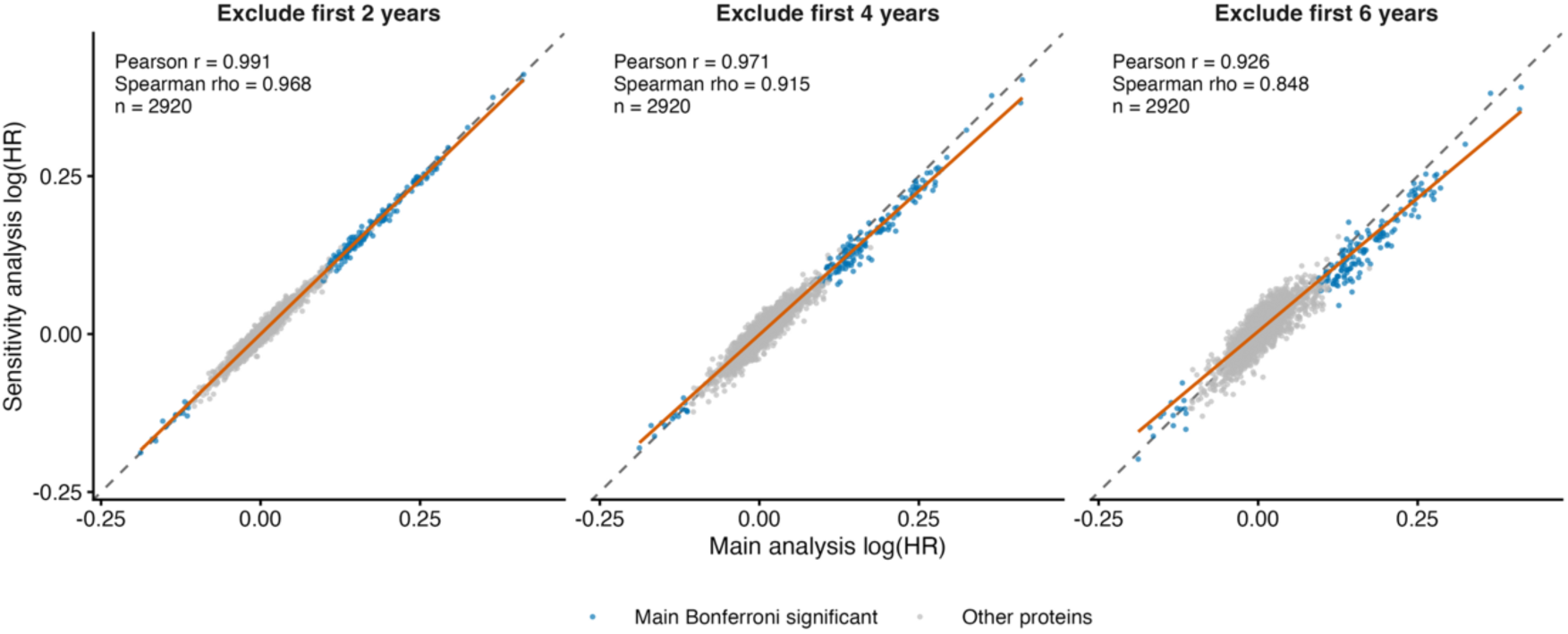
Lag sensitivity analyses excluding early incident COPD events. Protein-COPD Cox regression analyses were repeated after excluding participants who developed COPD within the first 2, 4, and 6 years of follow-up. Each panel compares protein log (HR) estimates from the lagged sensitivity analysis with those from the main training-set analysis. Pearson and Spearman correlations summarize the stability of protein effect estimates after reducing potential reverse causation.

**Fig. S6.**
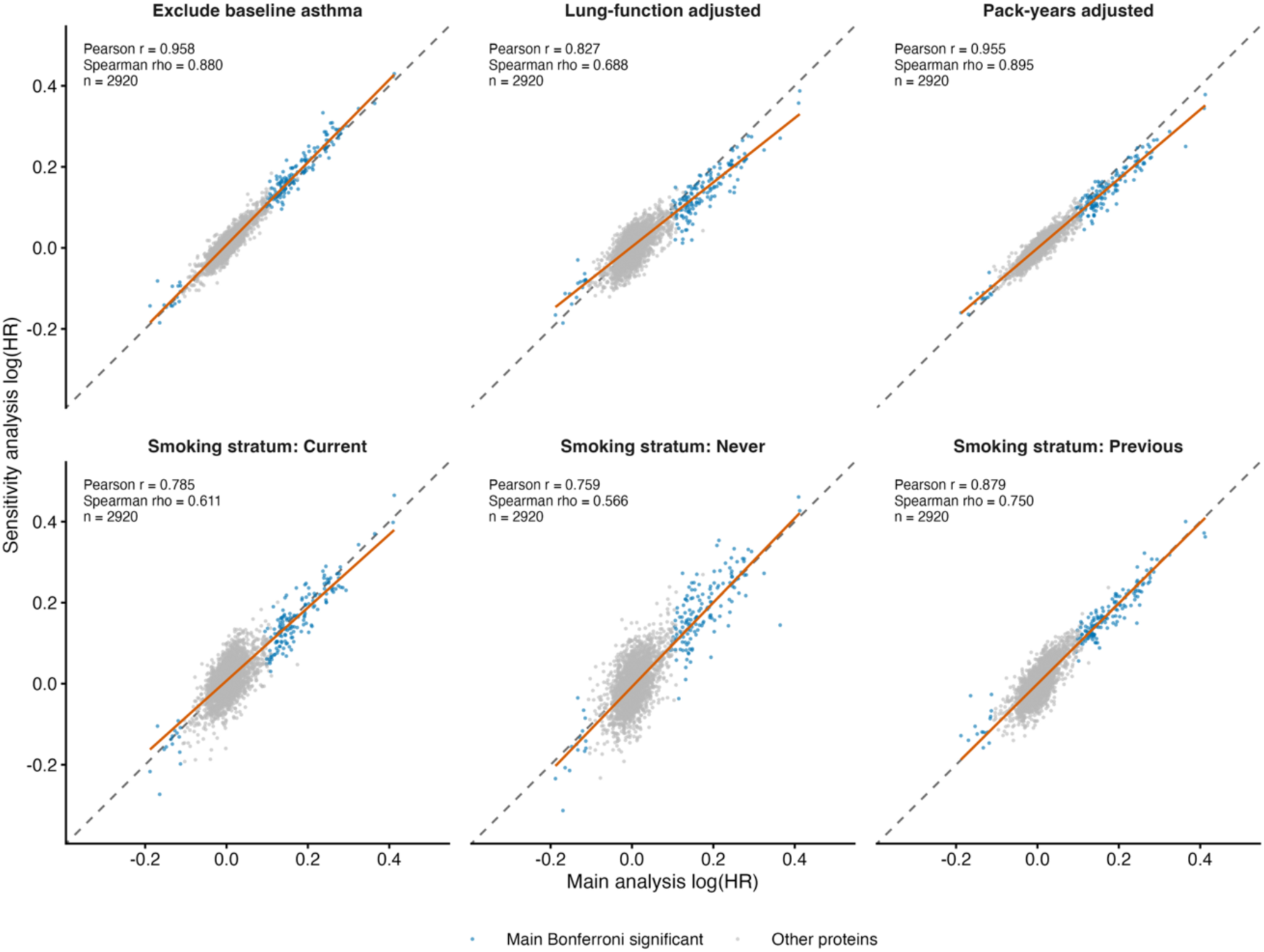
Additional sensitivity analyses for protein-COPD associations. Protein-wide Cox regression results from six sensitivity analyses were compared with the main training-set analysis: additional adjustment for smoking pack-years, additional adjustment for lung-function traits, exclusion of participants with baseline asthma, and analyses stratified by never, previous, and current smoking status. Each panel shows the correlation between main-analysis and sensitivity-analysis log (HR) estimates, with Bonferroni-significant proteins from the main analysis highlighted.

**Table S1.** Baseline characteristics of participants stratified by COPD incident in the training set.

|  | **Without COPD** | **With COPD** | **P** |
| --- | --- | --- | --- |
| N | 33422 | 1699 |  |
| Age (mean (SD)) | 56.47 (8.23) | 61.11 (6.54) | <0.001 |
| BMI (mean (SD)) | 27.38 (4.74) | 28.41 (5.56) | <0.001 |
| TDI (mean (SD)) | -1.30 (3.12) | -0.03 (3.55) | <0.001 |
| Sex (%) |  |  |  |
| Female | 18291 (54.7) | 774 (45.6) | <0.001 |
| Male | 15131 (45.3) | 925 (54.4) |  |
| Smoking status (%) |  |  |  |
| Never | 18924 (56.6) | 354 (20.8) | <0.001 |
| Previous | 11472 (34.3) | 748 (44.0) |  |
| Current | 3026 (9.1) | 597 (35.1) |  |
| Ethnic (%) |  |  |  |
| White | 31321 (93.7) | 1645 (96.8) | <0.001 |
| Others | 2101 (6.3) | 54 (3.2) |  |
| Education (%) |  |  |  |
| Low | 5454 (16.3) | 646 (38.0) | <0.001 |
| Medium | 7468 (22.3) | 354 (20.8) |  |
| High | 20500 (61.3) | 699 (41.1) |  |

**Table S2.** Baseline characteristics of participants stratified by COPD incident in the validation set.

|  | **Without COPD** | **With COPD** | **P** |
| --- | --- | --- | --- |
| N | 14323 | 728 |  |
| Age (mean (SD)) | 56.46 (8.21) | 61.08 (6.54) | <0.001 |
| BMI (mean (SD)) | 27.39 (4.68) | 28.45 (5.38) | <0.001 |
| TDI (mean (SD)) | -1.29 (3.12) | -0.23 (3.51) | <0.001 |
| Sex (%) |  |  |  |
| Female | 7785 (54.4) | 340 (46.7) | <0.001 |
| Male | 6538 (45.6) | 388 (53.3) |  |
| Smoking status (%) |  |  |  |
| Never | 8169 (57.0) | 170 (23.4) | <0.001 |
| Previous | 4869 (34.0) | 317 (43.5) |  |
| Current | 1285 (9.0) | 241 (33.1) |  |
| Ethnic (%) |  |  |  |
| White | 13424 (93.7) | 705 (96.8) | 0.001 |
| Others | 899 (6.3) | 23 (3.2) |  |
| Education (%) |  |  |  |
| Low | 2296 (16.0) | 292 (40.1) | <0.001 |
| Medium | 3157 (22.0) | 147 (20.2) |  |
| High | 8870 (61.9) | 289 (39.7) |  |

**Table S3.** Model metrics across three feature sets.

| **Feature** | **AUC (95% CI)** | **Accuracy** | **sensitivity** | **specificity** | **PPV** | **NPV** | **F1** | **Brier** |
| --- | --- | --- | --- | --- | --- | --- | --- | --- |
| protein + covariates | 0.824 [0.808-0.839] | 0.738 | 0.753 | 0.738 | 0.127 | 0.983 | 0.218 | 0.150 |
| protein | 0.804 [0.788-0.820] | 0.755 | 0.710 | 0.757 | 0.129 | 0.981 | 0.219 | 0.156 |
| covariates | 0.800 [0.784-0.816] | 0.701 | 0.749 | 0.699 | 0.112 | 0.982 | 0.195 | 0.178 |

**Notes:** AUC, area under the receiver operating characteristic curve; CI, confidence interval; PPV, positive predictive value; NPV, negative predictive value; F1, F1-score; Brier, Brier score. Accuracy, sensitivity, specificity, PPV, NPV, and F1-score are reported as proportions. Lower Brier scores indicate better calibration performance.

## Notes

### Competing Interest Statement

The authors have declared no competing interest.

### Author Declarations

The North West Multi-centre Research Ethics Committee gave ethical approval for the UK Biobank study (reference number 11/NW/0382). All participants provided written informed consent. This research was conducted using the UK Biobank Resource under Application Number 822932.

